# The effect of non-pharmaceutical interventions (NPIs) on the spread of COVID-19 pandemic in Japan: A modeling study

**DOI:** 10.1101/2020.05.22.20109660

**Authors:** Yingying Sun, Jikai Sun

**Author notes:** Correspondence: Jikai Sun, Department of Architecture and Architectural Engineering, Kyoto University, Gokasho, Uji, Kyoto, 6110011 Japan.

## Abstract

Non-pharmaceutical interventions (NPIs) are founded to be effective to delay epidemic spread and to reduce the number of patients. Moderate NPIs took in Japan seemed to have reduced the COVID-19 patients and to lower death rates, thus, effects of those NPIs are worthy of investigation. We used open source data and divided the data into three periods: Jan 22 to Feb 25 (Period I), Feb 26 to Apr 6 (Period II), and Apr 7 to May 14 (Period III). We developed the SIRD model and applied the Monte Carlo Simulation to estimate a combination of optimal results, including the peak of infected cases, the peak date, and *R*_0_. For Period I, the estimated peak infected cases were smaller than the observed ones, the peak date was earlier than the observed one, and the *R*_0_ was about 4.66. For the other two periods, the estimated cases were more, and the peak dates were earlier than the observed ones. The *R_0_* was 2.50 in Period II, and 1.79 in Period III. NPIs took in Japan might have reduced more than 50% of the daily contacts per people compared to that before COVID-19. Owing to the effects of NPIs, the Japanese society had avoided collapse of medical service. Nevertheless, the capacity of daily RT-PCR may have restricted the reported confirmed cases.

## 1. Introduction

Since December 2019, the novel coronavirus disease 2019 (COVID-19) epidemic caused by severe acute respiratory syndrome coronavirus 2 (SARS-CoV-2) have been reported in Wuhan, a city in the Chinese province of Hubei (China CDC, 2020). As of May 15, 2020, 4,338,658 confirmed cases, including 297,119 deaths, had been reported all over the world (WHO, 2020). In the absence of vaccines and available treatment, delaying virus spread and reducing the number of people infected by COVID-19 seemed only possible by implementing non-pharmaceutical interventions (NPIs) (Ferguson et al., 2020; Jester, Uyeki, & Jernigan, 2020; Lai et al., 2020; Wilder-Smith & Freedman, 2020). Up to now, a volume of studies have reported that, the lockdown of Wuhan was associated with a delayed arrival time of COVID-19 in other Chinese cities by an estimated 3 to 5 days (Chinazzi et al., 2020; Tian et al., 2020), and had a marked effect on the international scale, where case importations were reduced by nearly 80% until mid-February (Chinazzi et al., 2020). Also, social distancing measures and school closure were found to be sufficient to control COVID-19, to reduce peak incidence, and to delay the epidemic (Zhang et al., 2020). Nevertheless, the effects of NPIs on containing COVID-19 could vary across countries and societies, due to cultural, lawful, and societal challenges, as well as the timing and the extent of implementation (Blevins, Jalloh, & Robinson, 2019; Cauchemez, Valleron, Boelle, Flahault, & Ferguson, 2008; Glass, Glass, Beyeler, & Min, 2006; Mitchell et al., 2011).

Japan, one of the countries neighboring to China, had adopted traditional NPIs such as social distancing, travel restrictions, school and workplace closures, but did not enforce community lockdown, contact tracing by using technologies to invade individual information, or large-scale RT-PCR tests (Prime Minister of Japan and His Cabinet, 2020a). Instead, Japan took a series of unique strategies including “voluntary event cancellation, cluster tracing, and ask people to wait as least four days since symptom onset until contact a doctor” (Prime Minister of Japan and His Cabinet, 2020a). As of May 14, Japan has experienced a total of 16,079 COVID-19 patients, including 687 deaths (Japan Ministry of Health Labour and Welfare, 2020). The death rate in Japan is lower than that in China (5.59%) and Italy (14.12%) (JHU CSSE, 2020), although Japan has the highest rate of aging population as of 26.6% in the world (Japan Statistics Bureau, 2015). Studies reported that the elderly, people who have basic disease such as diabetes, cancers, or heart disease could be the most vulnerable groups to COVID-19 (Guan et al., 2020; Zhang et al., 2020; L. Zhang et al.). Therefore, effects of NPIs took by Japan are worthy of investigation.

## 2. Ethics statement

We used open data source to collect data. No ethical issue exists.

## 3. Data collection

Data of COVID-19 of Japan were retrieved from Center for Systems Science and Engineering, Johns Hopkins University (JHU CSSE, 2020). According to countermeasure policies for COVID-19 containment in Japan, we divided the data into three periods: Jan 22 to Feb 25 (Period I), Feb 26 to Apr 6 (Period II), and Apr 7 to May 14 (Period III). The Japanese government gradually enforced more rigorous countermeasure for COVID-19 containment through these three periods. For instance, on Feb 26, large-scale sports and entertainment events were canceled throughout Japan, and organizers of small-scale events were advised to voluntarily cancel events (Prime Minister of Japan and His Cabinet, 2020b). On Apr 7, emergency declaration was first declared to seven prefectures by Prime Minister of Japan, and were gradually declared to other prefectures (Figure 1) (Prime Minister of Japan and His Cabinet, 2020b).

**Figure 1.**
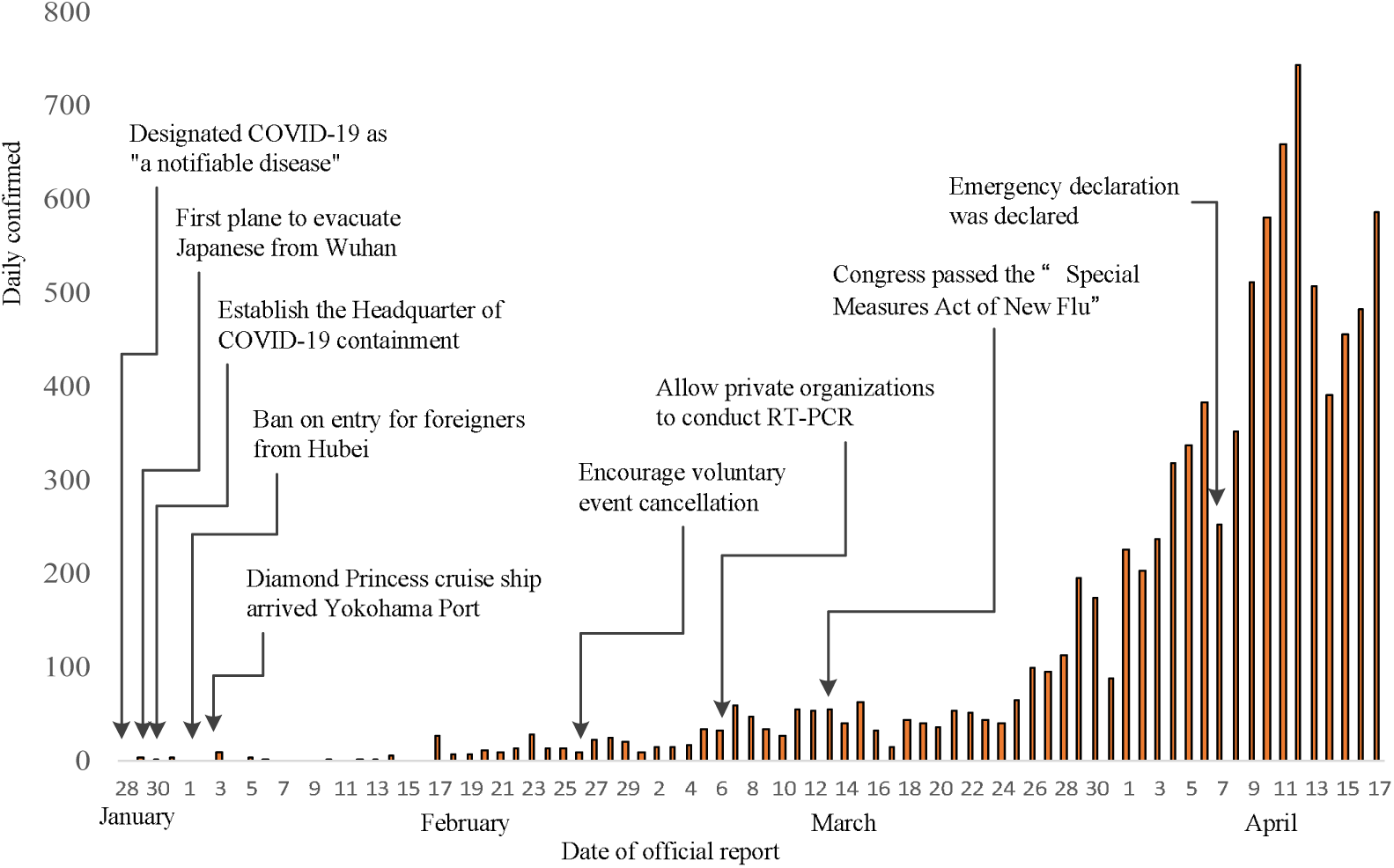
Timeline of countermeasures of COVID-19 in Japan

## 4. Method

To illustrate the effects of NPIs on the dynamics of the epidemic, we developed a simple Susceptible-Infectious-Recovered-Deceased (SIRD) Model (Osemwinyen & Diakhaby, 2015) of COVID-19 transmission. This model assumes that the total population of reference is constant. The population is divided into several different compartments (i.e., susceptible, infective, recovered, deceased) and specify how agents move across the separate compartments over time. The model assumes that susceptible individuals can become infectious after contact with an infectious individual, and recovered individuals cannot be re-infected. The SIRD model uses the following system of differential equations:

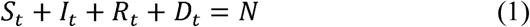

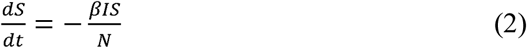

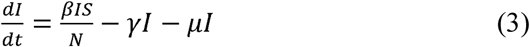

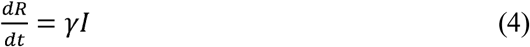

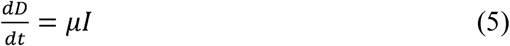

where *β*, *γ*, *μ* are the rates of infection, recovery, and mortality, respectively. In addition, a key parameter regulating the dynamics of an epidemic is the basic reproduction number (*R*_0_), which correspondes to the average number of secondary cases generated by an index case in a fully susceptible population (Zhang et al., 2020).

Using the SIRD model, we calculated the peak number of existing infected cases and the peak date. We compared the differences between estimated results and official reports, in order to estimate the effects of NPIs. According to the basis of plausible ranges from the recent reports of COVID-19 data (Bhatraju et al., 2020; Geleris et al., 2020; Guan et al., 2020; Li et al., 2020; Mehra, Desai, Kuy, Henry, & Patel, 2020; J. Zhang et al.), we consider that the average incubation period ranges from 2 to 14 days (*β* = 1/14, 1/2]), the recovery period ranges from 7 to 28 days (γ = [1/28 − 1/7]), and the deceased period ranges from 14 to 60 days (*μ* = [1/60 − 1/14]).

For the N, which is the total population of the people who may be exposed to the virus, was assumed to range between a baseline value and a maximum value. The baseline value equals to the number of people who received the reverse transcription polymerase chain reaction (RT-PCR) test in Japan. The maximum value equals to the product of “the baseline value * the average daily number of contacts per Japanese people”.

For Period I, no rigorous measures were taken to prevent COVID-19. Most of the laboratory-confirmed cases of COVID-19 were reported in large cities such as Tokyo, Yokohama, and Kyoto (Japan Ministry of Health Labour and Welfare, 2020). Thus, we assume that the daily contacts per people in Japan is 18.8, which is the same as that reported in Shanghai (Zhang et al., 2020). The total number of people who received RT-PCR was 1,846 until Feb 25 (Japan Ministry of Health Labour and Welfare, 2020). Hence, *N _I_* = [1846, 1846*18.8].

For Period II, voluntary event cancellation, school and workplace closures were carried out throughout Japan. The COVID-19 epidemic has already gradually spread to small cities and towns. Thus, we assume that the daily contacts per people in Japan is 18.8 * 0.7 * 0.5. The coefficient of “0.7” means that the Japanese daily contacts are about 70% of that in Shanghai, and “0.5” means that NPIs have reduced 50% of daily contacts. The total number of people who received RT-PCR was 46,172 until Apr 6 (Japan Ministry of Health Labour and Welfare, 2020). Hence, *N _II_* = [46172, 46172*18.8*0.7*0.5].

For Period III, emergency declaration was declared, and the Japanese government aimed to reduce the daily contacts per people to 20% compared to that before COVID-19 (Prime Minister of Japan and His Cabinet, 2020a). The total number of people who received RT-PCR was 233,144 until May 14 (Japan Ministry of Health Labour and Welfare, 2020). Hence, *N _III_* = [233144, 233144*18.8*0.7*0.2].

Based on the values of daily contacts per people in those three periods, we calculated that the average daily contacts per people throughout the entire period was 18.8*0.48. Hence, *N_Entire_* = [233144, 233144*18.8*0.48].

We coded the SIRD model into the Monte Carlo simulation, and applied machine learning to produce a bevy of results of, and N. We extracted the optimal combination of, and N from 10 groups of simulations. For each simulation, we produced 50,000 cases of the SIRD model. We wrote the code package and run the simulation by Python 3.8.

## 5. Results

### 5.1 The entire period (Jan 22 to May 14)

Based on the official reports (JHU CSSE, 2020), the distribution of total confirmed cases and the existing infected cases were shown in Figure 2. The optimal results of is 3.94, is 7.05, is 37.4, and N is 270,897. The is 1.79. Based on these optimal results, we produced the estimated number of existing infected cases, shown in Figure 3.

**Figure 2.**
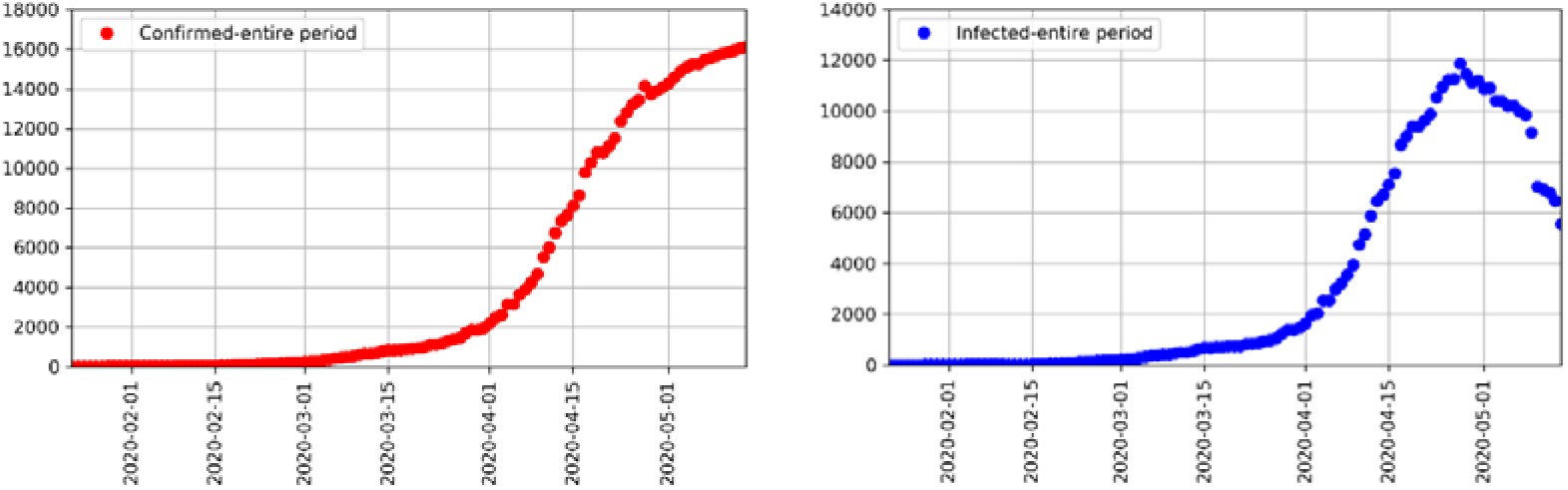
Confirmed cases and existing infected cases from Jan 22 to May 14.

**Figure 3.**
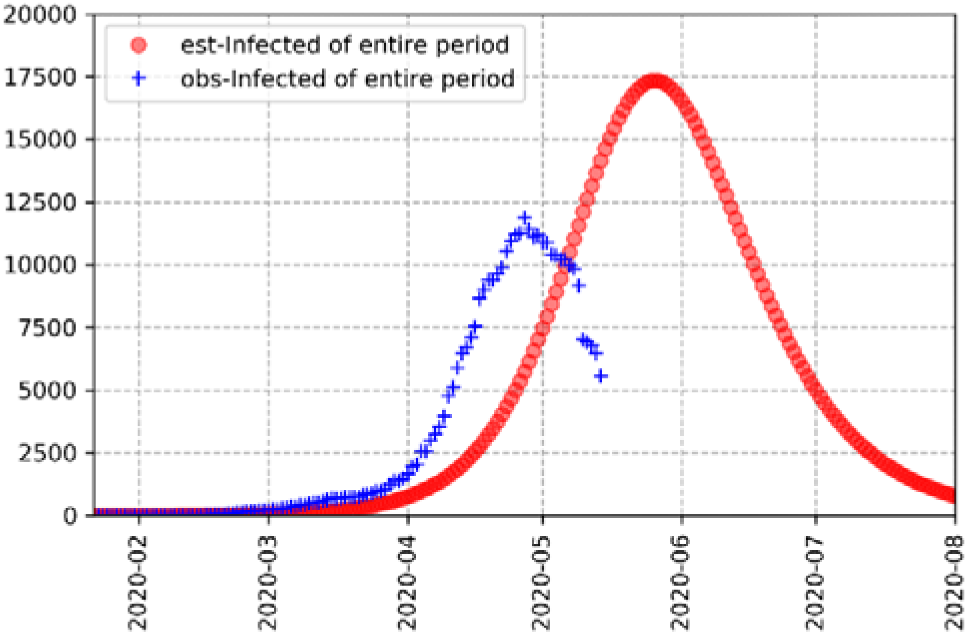
Estimated and observed numbers of existing infected cases from Jan 22 to May 14.

Our simulation results displayed that the estimated peak number of existing infected cases would be 17,344 on May 26. However, the official reports showed that the peak number of existing infected cases was 11,869 on April 27. These results indicated that 1) NPIs took in Japan might have reduced more than 52% of the average daily number of contacts per people compared to that before COVID-19; and 2) owing to the effects of NPIs, the Japanese society had avoided collapse of medical service. Nevertheless, 3) the capacity of daily RT-PCR may have restricted the reported confirmed cases; and 4) the official reports may have provided the underestimated figure of COVID-19.

### 5.2 Period I (Jan 22 to Feb 25)

The optimal results of is 5.41, is 25.22, is 53.66, and N is 34,347. The is 4.66. Based on these optimal results, we produced the estimated number of existing infected cases, shown in Figure 4.

**Figure 4.**
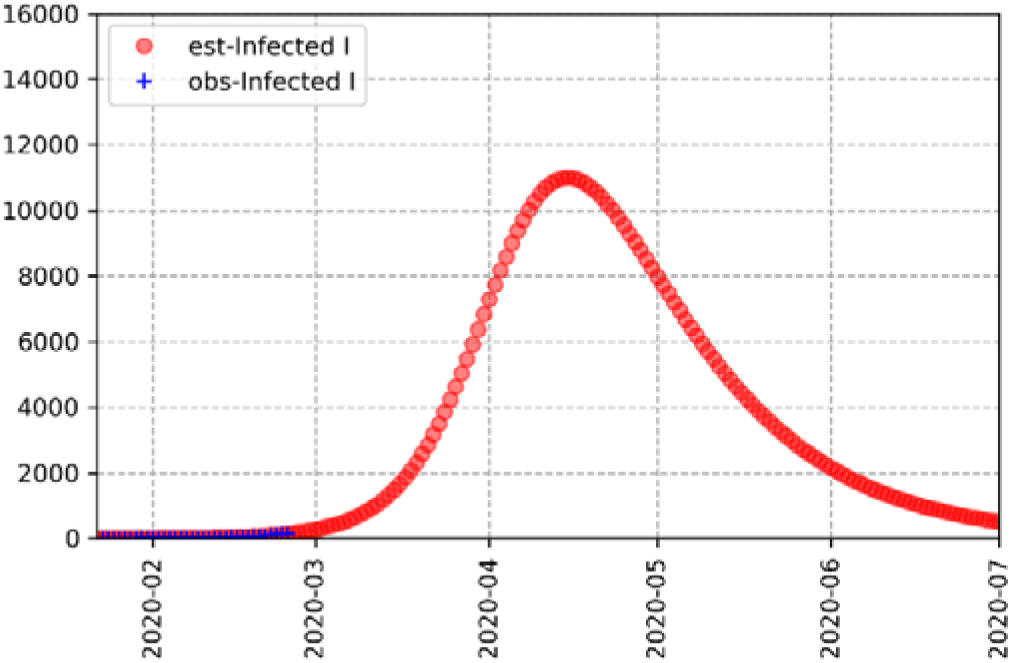
Estimated and observed numbers of existing infected cases from Jan 22 to Feb 25.

Our simulation results displayed that the estimated peak number of existing infected cases would be 11,010 on Apr 15. Compared to the official reports, which showed that the peak number of existing infected cases was 11,869 on April 27, these results indicated that 1) the capacity of daily RT-PCR obviously restricted the reported confirmed cases; 2) the of Period I was higher than that reported in Wuhan prior to the lockdown on 23 Jan (Tian et al., 2020); and 3) rigorous countermeasures were obviously required to contain COVID-19.

The optimal results of is 6.37, is 15.94, is 40.74, and N is 135,262. The is 2.50. Based on these optimal results, we produced the estimated number of existing infected cases, shown in Figure 5.

**Figure 5.**
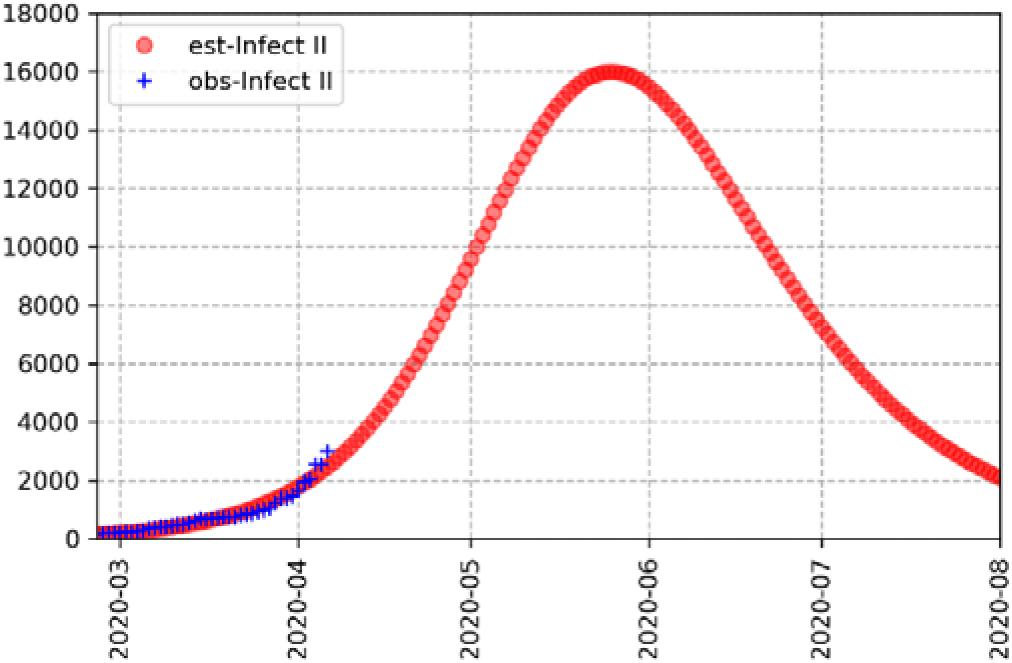
Estimated and observed number of existing infected cases from Feb 26 to Apr 6.

Our simulation results displayed that the estimated peak number of existing infected cases would be 15,992 on May 25. Compared to the official reports, which showed that the peak number of existing infected cases was 11,869 on April 27, these results indicated that 1) NPIs took in Period II might reduce more than 50% of the daily contacts per people compared to that before COVID-19; 2) owing to the effects of NPIs, the Japanese society had avoided collapse of medical service, which had been seen in Italy during the same period (Remuzzi & Remuzzi, 2020); 3) the of Period II was close to that reported in China before March 2020 (Chinazzi et al., 2020). Nevertheless, 4) the capacity of daily RT-PCR may have restricted the reported confirmed cases.

### 5.4 Period III (Apr 7 to May 14)

The optimal results of is 4.09, is 7.33, is 22.59, and N is 260,156. The is 1.79. Based on these optimal results, we produced the estimated number of existing infected cases, shown in Figure 6.

**Figure 6.**
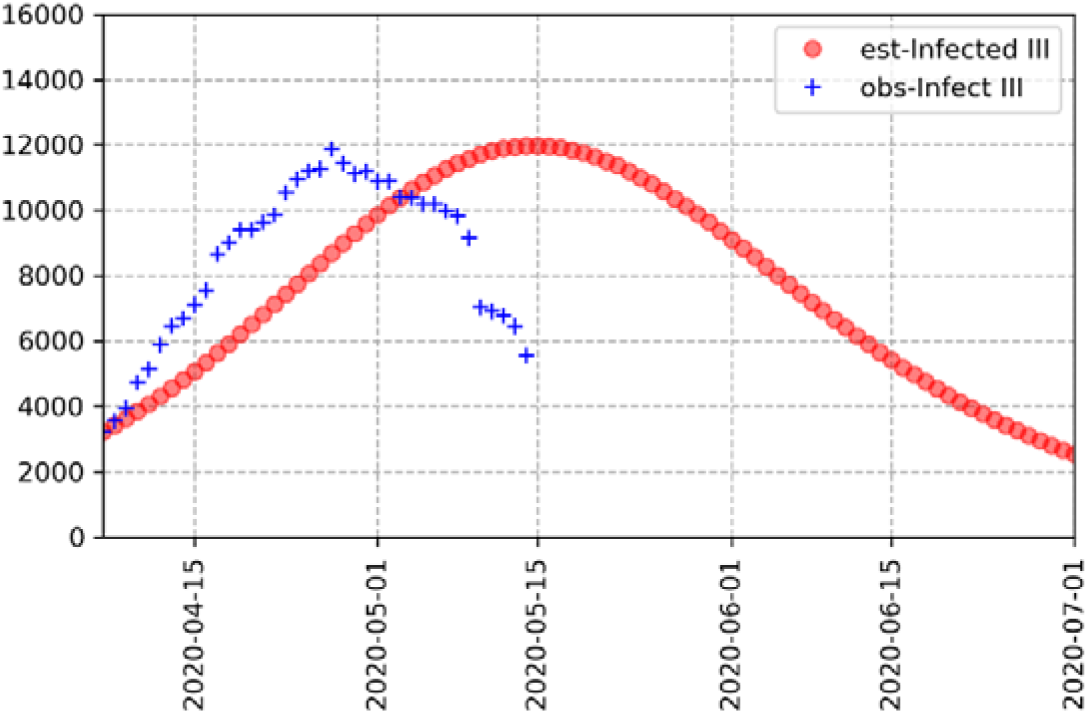
Estimated and observed numbers of existing infected cases from Apr 7 to May 14.

Our simulation results displayed that the estimated peak number of existing infected cases would be 11,972 on May 15. Compared to the official reports, which showed that the peak number of existing infected cases was 11,869 on April 27, these results indicated that 1) NPIs took in Period II might have reduced about 80% of the daily contacts per people compared to that before COVID-19; 2) owing to the effects of NPIs, the epidemic transmission may mainly have occurred through family contacts other than social contacts such as workplace (Zhang et al., 2020); and 3) the *R*_0_ of Period III was close to that reported throughout the entire period, which means that, the capacity of daily RT-PCR during Period III may have satisfied the required numbers.

## 6. Discussion

We developed an SIRD model to estimate the peak number of existing infected COVID-19 cases, the peak date, and the *R*_0_ in different periods. For an aging society like Japan, the most important prediction is the maximum number of patients who will require medical service. This prediction is of crucial importance to design for policies of COVID-19 containment in Japanese hospitals and to calculate the time period in which they need to be available. Meanwhile, as it is the first time to declare large-scale emergency declarations of infectious disease in Japan, the prediction is of fundamental significance to plan for social countermeasures such as NPIs.

For the Monte Carlo Simulation, instead of using the total population of Japan as the N population (Sugishita, Kurita, Sugawara, & Ohkusa, 2020), we used the number of people who received RT-PCR to estimate the developing trends of epidemic transmission. This method can generate more reliable predictions of the number of existing infected cases than using other methods. Reasons are that, first, we consider that the total confirmed cases are less challengeable for medical service than the existing infected cases. Studies have reported that, about 80% of COVID-19 patients did not need medical treatment, and thus do not exert any burdens on medical service (Prime Minister of Japan and His Cabinet, 2020a). Second, using the number of RT-PCR receivers to predict the peak incident can display the effectiveness of countermeasure policies. For instance, in Period II, the estimated peak incident is much more serious than the observed fact, meaning that the capacity of daily RT-PCR in Japan cannot satisfy the required numbers. Third, the prediction of peak incident in each period is crucial for the preparation of the minimum number of hospital beds. For instance, based on estimations of the peak incident by Japanese experts, the total hospital beds for COVID-19 patients have increased from about 2,000 in early February to 31,289 in May 2020 (Japan Ministry of Health Labour and Welfare, 2019).

Our simulation results showed that despite Period I, the estimated peak date could appear 18 to 29 days later than the observed date. The biggest reason might be because of the capability of daily RT-PCR, which directly defines the observed confirmed cases. Another reason might be the high level of Japanese medical service. On Jan 28, the Japanese government announced that the COVID-19 epidemic has been defined as “a designated notifiable disease” according to the Law of Infectious Disease and Patient Treatment, and medical treatment expenses of COVID-19 patients will be covered by the national budget (Prime Minister of Japan and His Cabinet, 2020b). Owing to stable and high-qualified medical service, the Japanese society seems to successfully have discharged COVID-19 patients and have avoided collapse of hospital service.

According to our simulation results, *R*_0_ in Period II is about 2.50, which is similar to that reported by previous studies in China and Italy (Chinazzi et al., 2020; Remuzzi & Remuzzi, 2020; Zhang et al., 2020). Nevertheless, *R*_0_ throughout the entire period was a smaller number as of 1.79. When considering that we used the number of RT-PCR receivers to produce the optimal results, the actual *R*_0_ might be even smaller than 1.79. This could be the reason why 39 Japanese prefectures have removed the emergency declaration of COVID-19 on Mar 14 (Prime Minister of Japan and His Cabinet, 2020a).

## 7. Conclusion

We applied machine learning to run the Monte Carlo Simulation, in order to produce the optimal results of estimated COVID-19 patients, the peak date, and the *R*_0_. According to the timing and the extent of NPIs implementation, we divided the data of Japanese COVID-19 epidemic into three periods. In this way, we were able to compare the estimated results with the observed reports, and to understand the effects of NPIs. Our results could contribute to the current COVID-19 studies by adding evidence on detailed information from Japan. But more significantly, our results provided empirical findings on the effects of NPIs implementation. These findings could help policy makers to plan for hospital beds, to establish strategies of NPIs, and to consider risk communication methods to the public.

Nevertheless, the study has a few limitations. Although we revealed a combination of COVID-19 transmission dynamics, we could not investigate further to the prefectural level. For instance, the total hospital beds for COVID-19 is much higher than the estimated and the observed existing patients, but the hospital admission capability vary across different prefectures. Future studies are needed to work on unveiling the detailed status of each prefecture. Also, the ranges of the N population could largely define the optimal results. Despite using the number of RT-PCR receivers, future researchers need to try to find other ways to enhance the accuracy of the N population.

## Data Availability

Data were retrieved from Center for Systems Science and Engineering, Johns Hopkins University.

## Author contribution

Yingying Sun designed the study and drafted the manuscript. Jikai Sun conducted modelling and simulation.

